# Justice and fairness in research priority setting exercises on climate and health: protocol for a systematic review

**DOI:** 10.1101/2025.02.28.25322976

**Authors:** Susan Joy Lake, Soumyadeep Bhaumik

## Abstract

Research priority setting (RPS) exercises are a collective activity, involving various interest-holders to reach consensus on what research should be prioritised. It enables research being more relevant to interest holders. With climate change being visible there is increasing research on climate and health. Consequently, research priority setting on climate and health domain has also become more common. RPS exercises can serve to sustain or enhance various types of injustices, be it epistemic injustice, climate injustices or other forms of injustice and unfair knowledge practices. On the other hand, it can also serve as pro-justice, pro-fairness tool in the knowledge ecosystem of climate and health. To understand how RPS exercises on climate and health consider, and address justice and fairness by systematically reviewing them.

## Introduction

Climate change has wide-reaching implications not only for humanity but also the biosphere and all life which depends upon it. It has been earmarked as the greatest threat to global health.^1,2^ These impacts include increase in extreme weather events; loss of species diversity; pollution of land, air and waterways; changes in infectious disease transmission; food and water insecurity; human displacement and widening health inequities.^1,3^ The threats of climate change are not distributed evenly across all people, as those who least contribute to climate change are affected the soonest and with most severe outcomes.^4^ With the threat of climate change being realised in reality, there is increasing research in the climate and health ecosystem, and consequently the need to set research priorities.

Research priority setting (RPS) exercises are a collective activity which brings together a diverse range of interest-holders in a formalised process to reach consensus on what research should be prioritised.^5^ RPS is essential as the resources with which to conduct research are finite and interest-holders benefit differentially depending on how resources are allocated. ^5^ Within the climate and health research context, RPS exercises can serve to sustain or enhance various types of injustices, be it epistemic injustice, climate injustices or other forms of injustice and unfair knowledge practices.^6-10^ On the other hand, it can also serve as pro-justice, pro-fairness tool in the knowledge ecosystem of climate and health.

## Objective

To understand how RPS exercises on climate and health consider, and address justice and fairness.

## Methods

### Registration and protocol

The protocol for this study is being registered a priori and the study will be reported as per PRISMA reporting guidelines. ^11^

### Ethics

This is a systematic review of existing literature. It does not involve any living participant.

### Eligibility criteria

We will include any priority setting exercises, using any method (formal or informal) which are explicitly conducted to set priorities for research on climate and health. We will take an inclusive approach so as to define what research on climate and health means. We will include RPS

- setting priorities for health research on climate change or due to extreme weather events
- setting priorities for health research on any health condition/topic but with recognition of the inter-relationship between health of humans, and climate or environment (with or without specific mention of frameworks like One Health, EcoHealth, Planetary health).

RPS will be included if they are published on or after 2015, irrespective of the geographical scope, intended beneficiaries, target audience of the priorities, health area, type of questions/topics prioritised, or time frame. Commentaries or perspective pieces outlining research priorities without any process at all will not be included.

### Information sources & Search Strategy

We will search the following bibliographic database:

1. PubMed
2. EMBASE
3. CINAHL
4. GreenFile
5. Family and Society Studies Worlwide

We will also search the following repositories to identify any RPS which might have not been published in journals:

- Repository of research priority setting by WHO
- LBG-OIS Centre Research Priority Setting Database

In addition, we will manually screen the reference list of RPS found by other search methods to identify additional RPS which might meet eligibility criteria and reach out to experts . Full search strategies of all bibliographic databases are mentioned in Supplementary Appendix.

### Selection process

At least two reviewers will independently screen all retrieved records, first at tittle -abstract level and then at full -text review level. Any disagreement will be resolved through team discussions .

### Data collection

Two reviewers will independently extract data using a standardised data extraction form through redcap, which has the following components:

i. Key Characteristics of the included RPS
ii. Procedural Justice in RPS
iii. Hermeneutical Justice in RPS
iv. Testimonial Justice in RPS
v. Aspects of justice specific to climate and health
vi. Fairness of RPS outcomes

These domains were identified to align with justice and fairness concepts. These concepts were then mapped to specific steps of the RPS, using the REPRISE reporting guideline^12^ as a base and then iteratively translating concepts to real-world aspects of the RPS. The data extraction form will be tested, and any changes will be built in.

Any discrepancies were resolved through consensus through team discussions.

### Synthesis methods

We will use a structured synthesis in alignment with different justice parameters. For numerically extracted data we will present data descriptively using percentages, proportions or rations as relevant. For data which is captured through open questions textually we will analyse data narratively.

## Data Availability

This is a protocol and all relevant data in relation to the protocol arec contained in the manuscript.

