## Supplementary Appendix for "Justice and fairness in research priority setting exercises on climate and health: protocol for a systematic review"

### Supplementary Appendix : Search Strategies

#### *1.1 Search strategy for PubMed*

1. "Health Priorities"[Mesh] AND research* [tiab]
2. "research priority" [tiab:~3]
3. "research priorities" [tiab:~3]
4. "research agenda" [tiab:~3]
5. “priority-setting research” [tiab:~3]
6. “agenda-setting research” [tiab:~3]
7. #1 OR #2 OR #3 OR #4 OR #5 OR #6
8. "Climate Change"[MeSH] OR "One Health"[Mesh] OR "Environmental Health"[Mesh]
9. (("Climate"[Mesh] OR "Environment"[Mesh] OR "Extreme Weather"[Mesh]) AND "Health" [Mesh])
10. ("climate"[ti] OR "environment*"[ti] OR "global warming"[ti] OR "carbon footprint"[ti] OR "decarbonisation"[ti] OR "sustainability"[ti] OR "net-zero" [ti] OR "planetary health"[ti] OR Ecohealth[ti] OR "heat wave"[ti] OR "heat-wave"[ti] OR "tornado*"[ti] OR “cyclone*”[ti] OR "tsunami*"[ti] OR flood*[ti] OR "sea level rise"[ti] OR "coastal erosion"[ti] OR "landslide*"[ti] OR "drought*"[ti] OR "wild fire"[ti] OR "bush fire*"[ti] OR "forest fire*"[ti] OR "cold wave*"[ti])
11. #8 OR #9 OR #10
12. #7 AND #11

Filters applied: from 2015/1/1 - 3000/12/12.

#### *1.2. Search strategy for Ovid Embase Classic+Embase <1947 to 2025 February*

1. exp research priority/
2. research priorit$.ab,ti.
3. research agenda.ab,ti.
4. "research priority-setting".ab,ti.
5. "research agenda-setting".ab,ti.
6. "priority setting for research".ab,ti.
7. "setting priorit$ for research".ab,ti.
8. 1 or 2 or 3 or 4 or 5 or 6 or 7
9. exp "Climate Change"/ OR exp "One Health"/ OR exp "Environmental Health"/
10. ((exp Climate/ OR exp Environment/ OR exp "Extreme Weather"/) AND exp Health/)
11. climate.ti. OR environment*.ti. OR "global warming".ti. OR "carbon footprint".ti. OR decarbonisation.ti. OR sustainability.ti. OR net-zero.ti. OR "planetary health".ti. OR Ecohealth.ti. OR heat-wave.ti. OR heat-wave.ti. OR tornado*.ti. OR cyclone*.ti. OR tsunami*.ti. OR flood*.ti. OR "sea level rise".ti. OR "coastal erosion".ti. OR landslide*.ti. OR drought*.ti. OR "wild fire".ti. OR "bush fire*".ti. OR "forest fire*".ti. OR "cold wave*".ti.
12. limit 11 to ("remove medline records" and yr="2015 -Current")

#### *1.3. Search strategy for CINAHL*

1. (((MH "Health Priorities+") AND (TI research* OR AB research*)) OR (TI "research priority" OR "research priorities" OR "research agenda" OR “priority-setting research" OR “research priority-setting" "research agenda-setting" OR "priority setting for research" OR "setting priorit$ for research") OR (AB "research priority" OR "research priorities" OR "research agenda" OR “priority-setting research" OR “research priority-setting" "research agenda-setting" OR "priority setting for research" OR "setting priorit$ for research"))
2. (MH "Climate Change+") OR (MH "One Health+") OR (MH "Environmental Health+")
3. (((MH Climate+) OR (MH Environment+) OR (MH "Extreme Weather+")) AND (MH Health+))
4. (TI climate) OR (TI environment*) OR (TI "global warming") OR (TI "carbon footprint") OR (TI decarbonisation) OR (TI sustainability) OR (TI net-zero) OR (TI "planetary health") OR (TI Ecohealth) OR (TI heat-wave) OR (TI heat-wave) OR (TI tornado*) OR (TI cyclone*) OR (TI tsunami*) OR (TI flood*) OR (TI "sea level

rise") OR (TI "coastal erosion") OR (TI landslide*) OR (TI drought*) OR (TI "wild fire") OR (TI "bush fire*") OR (TI "forest fire*") OR (TI "cold wave*")
5. S2 OR S3 OR S4
6. S1 AND S5

Filters applied

- Exclude MEDLINE records

Expanders

- Apply equivalent subjects

Limiters

- Date Published: 20150101-

#### *1.4. Search strategy for Family & Society Studies Worldwide*

1. (((MH "Health Priorities+") AND (TI research* OR AB research*)) OR (TI "research priority" OR "research priorities" OR "research agenda" OR "priority-setting research" OR "research priority-setting" "research agenda-setting" OR "priority setting for research" OR "setting priorit$ for research") OR (AB "research priority" OR "research priorities" OR "research agenda" OR "priority-setting research" OR "research priority-setting" "research agenda-setting" OR "priority setting for research" OR "setting priorit$ for research"))
2. (MH "Climate Change+") OR (MH "One Health+") OR (MH "Environmental Health+")
3. (((MH Climate+) OR (MH Environment+) OR (MH "Extreme Weather+")) AND (MH Health+))
4. (TI climate) OR (TI environment*) OR (TI "global warming") OR (TI "carbon footprint") OR (TI decarbonisation) OR (TI sustainability) OR (TI net-zero) OR (TI "planetary health") OR (TI Ecohealth) OR (TI heat-wave) OR (TI heat-wave) OR (TI tornado*) OR (TI cyclone*) OR (TI tsunami*) OR (TI flood*) OR (TI "sea level rise") OR (TI "coastal erosion") OR (TI landslide*) OR (TI drought*) OR (TI "wild fire") OR (TI "bush fire*") OR (TI "forest fire*") OR (TI "cold wave*")
5. S2 OR S3 OR S4
6. S1 AND S5

Filters applied

- Exclude MEDLINE records

Expanders

- Apply equivalent subjects

Limiters

- Date Published: 20150101-

#### ***1.5.*** *Search strategy for GreenFile*

1. (((MH "Health Priorities+") AND (TI research* OR AB research*)) OR (TI "research priority" OR "research priorities" OR "research agenda" OR "priority-setting research" OR "research priority-setting" "research agenda-setting" OR "priority setting for research" OR "setting priorit$ for research") OR (AB "research priority" OR "research priorities" OR "research agenda" OR "priority-setting research" OR "research priority-setting" "research agenda-setting" OR "priority setting for research" OR "setting priorit$ for research"))
2. (MH "Climate Change+") OR (MH "One Health+") OR (MH "Environmental Health+")
3. (((MH Climate+) OR (MH Environment+) OR (MH "Extreme Weather+")) AND (MH Health+))

4. (TI climate) OR (TI environment*) OR (TI "global warming") OR (TI "carbon footprint") OR (TI decarbonisation) OR (TI sustainability) OR (TI net-zero) OR (TI "planetary health") OR (TI Ecohealth) OR (TI heat-wave) OR (TI heat-wave) OR (TI tornado*) OR (TI cyclone*) OR (TI tsunami*) OR (TI flood*) OR (TI "sea level rise") OR (TI "coastal erosion") OR (TI landslide*) OR (TI drought*) OR (TI "wild fire") OR (TI "bush fire*") OR (TI "forest fire*") OR (TI "cold wave*")
5. S2 OR S3 OR S4
6. S1 AND S5

Expanders

- Apply equivalent subjects

Limiters

- Date Published: 20150101-
